# A small number of early introductions seeded widespread transmission of SARS-CoV-2 in Québec, Canada

**DOI:** 10.1101/2021.03.20.21253835

**Authors:** Carmen Lía Murall, Eric Fournier, Jose Hector Galvez, Arnaud N’Guessan, Sarah J. Reiling, Pierre-Olivier Quirion, Sana Naderi, Anne-Marie Roy, Shu-Huang Chen, Paul Stretenowich, Mathieu Bourgey, David Bujold, Romain Gregoire, Pierre Lepage, Janick St-Cyr, Patrick Willet, Réjean Dion, Hugues Charest, Mark Lathrop, Michel Roger, Guillaume Bourque, Jiannis Ragoussis, B. Jesse Shapiro, Sandrine Moreira, on behalf of the Coronavirus Sequencing in Québec (CoVSeQ) Consortium

## Abstract

Using genomic epidemiology, we investigated the arrival of SARS-CoV-2 to Québec, the Canadian province most impacted by COVID-19, with >280,000 positive cases and >10,000 deaths in a population of 8.5 million as of March 1^st^, 2021. We report 2,921 high-quality SARS-CoV-2 genomes in the context of >12,000 publicly available genomes sampled globally over the first pandemic wave (up to June 1^st^, 2020). By combining phylogenetic and phylodynamic analyses with epidemiological data, we quantify the number of introduction events into Québec, identify their origins, and characterize the spatio-temporal spread of the virus. Conservatively, we estimated at least 500 independent introduction events, the majority of which happened from spring break until two weeks after the Canadian border closed for non-essential travel. Subsequent mass repatriations did not generate large transmission lineages (>50 cases), likely due to mandatory quarantine measures in place at the time. Consistent with common spring break and ‘snowbird’ destinations, most of the introductions were inferred to have originated from Europe via the Americas. Fewer than 100 viral introductions arrived during spring break, of which 5-10 led to the largest transmission lineages of the first wave (accounting for 36-58% of all sequenced infections). These successful viral transmission lineages dispersed widely across the province, consistent with founder effects and superspreading dynamics. Transmission lineage size was greatly reduced after March 11^th^, when a quarantine order for returning travelers was enacted. While this suggests the effectiveness of early public health measures, the biggest transmission lineages had already been ignited prior to this order. Combined, our results reinforce how, in the absence of tight travel restrictions or quarantine measures, fewer than 100 viral introductions in a week can ensure the establishment of extended transmission chains.

## Introduction

Over a year into the SARS-CoV-2 pandemic, whole-genome sequencing combined with phylogenetic analysis has emerged as an essential tool to track the local and global spread and evolution of the virus (Martin, VanInsberghe, and Koelle 2021; du Plessis et al. 2021). While the pandemic is global by definition, regional instances of viral introductions and spread provide ‘natural experiments’ to gain insights into general patterns. For example, Russia, Scotland and Massachusetts all experienced dozens to a few hundred independent introduction events of the virus from different locations (Komissarov et al. 2021; da Silva Filipe et al. 2021; Lemieux et al. 2021). The phylogenetic analysis of Massachusetts in particular found that most introduced viruses went extinct, while a minority of introductions were highly successful, consistent with superspreading dynamics (Lemieux et al. 2021). Phylogenetic analysis can also identify cryptic transmission chains unidentified by contact tracing or travel history (Bedford et al. 2020; Worobey et al. 2020). More recently, evidence has accumulated that transmissibility can be increased by adaptive mutations in the viral genome, such as amino acid change D614G in the spike protein (Volz et al. 2021; Zhou et al. 2021), or combinations of mutations, as in the B.1.1.7 lineage that emerged in Southeast England in September 2020 and is now the predominant lineage in the United Kingdom (Davies et al. 2021). The interplay between adaptive evolution, such as beneficial mutations, and stochastic factors, such as founder effects and superspreading, remains to be fully explored, and additional case studies are instructive to distinguish region-specific from generalizable features of the pandemic.

The province of Québec (QC) was the epicenter of the first wave in Canada of the SARS-CoV-2 pandemic (defined here up to June 1^st^, 2020). It is the second most populous province, with about half of its 8.5 million inhabitants in the densely populated Montréal metropolitan area. By June 1^st^, 2020, 5,210 people in Québec had died of Coronavirus disease 2019 (COVID-19), of whom 70% were residents of long-term care facilities. When the first cases were reported in China and Europe, the Public Health Laboratory of Québec (LSPQ) developed a qPCR diagnostic test targeting SARS-CoV-2 E and N genes (LeBlanc et al. 2020). The first case of COVID-19 in Québec was detected on February 25^th^, 2020. Shortly after, Québec was the first large Canadian province to start its spring school holiday (“spring break;” February 29^th^ to March 9^th^, 2020; **Fig. 1A**). It is believed that international travellers returning from spring break had a large impact on the epidemic (Godin et al. 2021). The number of cases increased exponentially during March 2020 (**Fig. 1A**; “Données COVID-19 Au Québec,” 2021). On March 13^th^, a public health emergency was declared, with schools, daycares and most other public spaces closed on March 16^th^ (“lockdown”). The closure of the Canadian border to non-essential travel was also announced March 16^th^ and officially closed the night of the 17^th^, except for returning Canadian citizens who continued to enter the country after repatriation calls from the government. On March 20^th^, Québec reached the threshold of 100 cases per day and by March 28^th^ random road checks were set up to discourage movement between regions within Québec and between neighbouring provinces (*i*.*e*., movement between Gatineau, Québec, bordering Ottawa, Ontario, was restricted). In April 2020, the virus spread significantly in long-term care facilities overwhelming many of them, thus requiring redeployment of health care workers and by April 20^th^ the Canadian Armed Forces sent personnel to the Montréal region to help. Having flattened the epidemic curve and with cases declining, public health measures began easing mid-May (**Fig.1A**). A year later, as of March 1^st^, 2021, Québec had suffered the highest death toll in Canada (over 10,000 dead) and among the highest death rates in the world (∼92 deaths per 100,000).

**Fig. 1.**
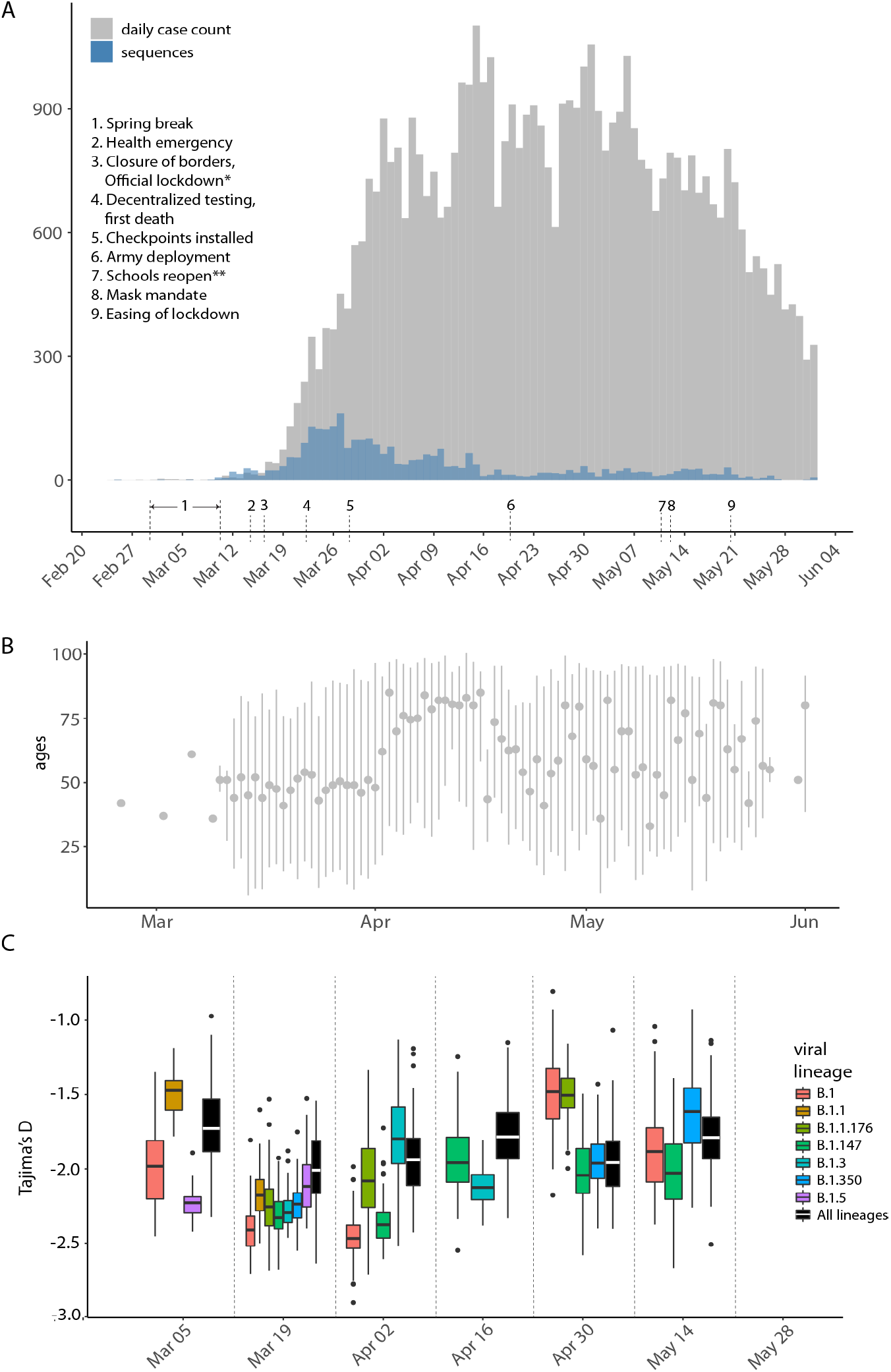
Timeline of COVID-19 cases and sequencing in Québec up to June 1. (A) Comparison of confirmed cases (grey) reported by public health authorities and high-quality sequences used in this study (blue) distributed by collection date. *Official lockdown included stay home orders and closure of schools and daycares. **Except schools in the city of Montreal. Timeline and control measures are from: (Rowe 2020; “Ligne Du Temps COVID-19 Au Québec” 2020). (B) Age distribution of sequenced cases (mean and range shown). (C) Variation in viral epidemiological dynamics as estimated by Tajima’s *D*. Boxplots represent 99 resampled estimates of Tajima’s *D* from random resamplings of 20 genome sequences for each two-week time period. Tajima’s *D* values are only estimated for Pangolin viral lineages with at least 20 sequences in a given time period.

In April 2020, we assembled the Coronavirus Sequencing in Québec (CoVSeQ) consortium of academic and government scientists (https://covseq.ca/) to sequence SARS-CoV-2 genomes in Québec. The CoVSeQ consortium is part of the Canadian COVID Genomic Network (CanCOGeN), a pan-Canadian cross-agency network for large-scale SARS-CoV-2 and human host sequencing (https://www.genomecanada.ca/en/cancogen). To better understand the early introductions and spread of SARS-CoV-2 in Québec during the first wave, we sequenced and analyzed 2,921 high-quality consensus genome sequences obtained between mid-February and June 1st, 2020. We studied how these Québec sequences were related to 12,801 genomes sampled from elsewhere in Canada and internationally. We inferred geographical origins of introduction events by comparing travel history data with phylogenetic inference, and estimated their likely arrival dates and subsequent spread. We conservatively estimated >500 independent introduction events, mainly involving viral lineages of European origin, of which the most successful arrived during the spring break period. Similarly to Massachusetts (Lemieux et al. 2021), viral transmission was overdispersed and suggestive of superspreading, with most lineages going extinct and only 5-10 introduction events giving rise to 50 or more cases. Consistent with founder effects and the effectiveness of public health measures, earlier introductions tended to give rise to more subsequent infections.

## Results and Discussion

### Sampling and sequencing the first wave in Québec

Our province-wide sequencing effort covers the first pandemic wave up to June 1^st^, 2020, with a focus on the earliest confirmed cases up to April 1^st^ (**Fig. 1A**). Consensus sequences of SARS-CoV-2 viral genomes were obtained by targeted amplification from clinical nasopharyngeal swabs specimens followed by sequencing on Nanopore (n=180), Illumina (n=2630) or MGI (n=111) platforms. Only sequences passing quality criteria (less than 5% undetermined bases, “Ns”) were considered for further phylogenetic analyses (Methods; **Table S1**). With these genome sequences we covered 5.7% of the total number of reported cases (45,641 laboratory confirmed cases and 5,849 suspected cases) up to and including June 1^st^. To capture early introduction events, our sequencing effort was highest (covering 27% of cases) before April 1^st^, two weeks after the Canadian border closed and most repatriation of Canadian citizens from abroad had occurred. Until early April, the mean age of sequenced cases was approximately 50 years old, then jumped to ∼75 years old, likely reflecting that the virus had entered long-term care facilities (**Fig. 1B**). By April 1^st^, over 500 long-term care facilities had reported at least one case of COVID-19, and the virus spread steadily through these primarily elderly populations during the month of April (Shingler 2020).

### Inferred SARS-CoV-2 introductions to Québec are mostly of European origin

Before federally mandated quarantine orders for returning travelers were put in place on March 25^th^, 1,544 travelers who entered Québec had tested positive with COVID-19 (Godin et al. 2021). However, not all these cases were necessarily independent introduction events, nor would they all give rise to successful onward transmission of SARS-CoV-2. To complement and refine the identification of introduction events, we compared self-reported travel history provided by COVID-19 positive cases with a phylogenetic inference of Québec and global context sequences (n = 15,722 viral genomes in total). Following previous phylogenetic studies of SARS-CoV-2 (du Plessis et al. 2021; Lemieux et al. 2021), we used ancestral state reconstruction (ASR) to identify non-Québec to Québec transition nodes in the phylogeny (Methods). In this way, we inferred a total of 555 (based on parsimony ASR) to 614 independent introduction events (based on maximum likelihood ASR, to which we refer below unless otherwise indicated). In our preliminary study of 734 Québec sequences up to April 1^st^, we estimated only 247 introduction events (“Genomic Epidemiology of Early Introductions of SARS-CoV-2 into the Canadian Province of Québec” 2020), suggesting that introductions are underestimated and are likely to increase with sample size. We defined Québec transmission lineages as descendants of a unique introduction event in the phylogeny, and then annotated these based upon Pangolin and Nextstrain lineage nomenclatures. Note that Pangolin or Nextstrain are viral phylogenetic lineages used for taxonomic purposes, which are distinct from Québec transmission lineages, which we define at higher phylogenetic resolution as descendants of a single introduction event, and thus represent a partially observed transmission chain.

We calculated Tajima’s *D* as a simple non-parametric metric of viral effective population size (Kim, Omori, and Ito 2017) and found strongly negative values of *D* early in the epidemic, consistent with exponential growth in mid-March to early April, followed by decelerating growth as public health measures likely reduced viral transmission (**Fig. 1C**). The decline of Tajima’s *D* from March 5 to 19th coincides with, or slightly precedes the increase in the epidemic curve starting on March 19th, suggesting its utility as an early indicator of population expansion. For example, viral lineage B.1, which originated in Italy and spread throughout Europe, showed evidence of rapid growth in Québec (median *D* ∼ –2.5 in mid-March) followed by a decline in late April and May. This is consistent with our observation that B.1 became very common in Québec by April before being replaced by other B.1 variants (notably B.1.147 and B.1.350) by the end of May (**Fig. S1**).

Of the 2,921 Québec consensus sequences analyzed here, 328 were from COVID-19 cases that had reported recent travel history. Note that a lack of travel history could indicate a true lack of travel, or a lack of available data. Travelers reported returning from the Caribbean and Latin America (n = 105, 32%, mainly from Mexico, n = 31, 9.5% and the Dominican Republic, n = 30, 9%), Europe (n = 104, 32% with the most from France, n = 39, 12% and Spain, n = 20, 6%), and the USA (n = 77, 24%) (**Fig. 2A**). There was very little reported travel from Asia (n = 4, 1.2%) and none from China. Only 153-162 phylogenetically inferred introduction events (25-28%; range across parsimony or ML methods) were associated with travel history. These events were broadly concordant with travel history, with some exceptions: notably, Latin America and Europe were approximately equally popular destinations based on travel history, but phylogenetic analysis identified Europe as the more likely origin of introductions into Québec (**Fig. 2A,B**). This is consistent with European viral lineages arriving in Québec, perhaps via the Americas – but before accumulating lineage-defining mutations in the Americas. The early introductions of viral lineages A and B.4, common in the early outbreaks in China and Iran respectively, appear not to have been successful in Québec and were not observed by mid-May (**Fig. S1**).

**Fig. 2:**
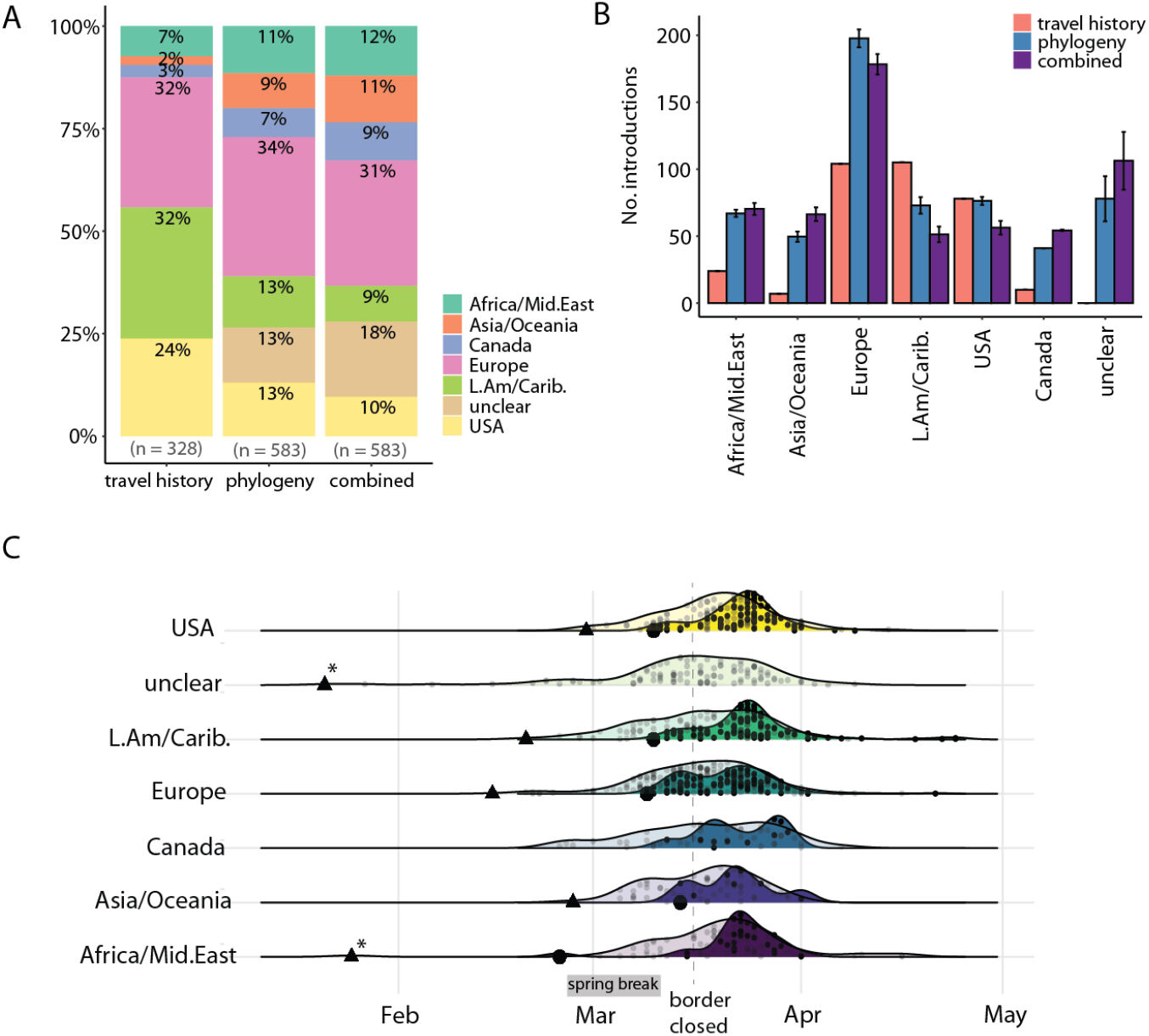
Analysis of introduction events. (A) Introduction event origins as a percentage of the total inferred by each method. (B) Number of introduction events by region of origin inferred by self-reported travel history, phylogenetic ancestral state reconstruction, or both combined. Bars denote the mean value with error bars indicating standard devotion (SD) across three ASR methods (ML, Deltran, Acctran). “Canada’’ refers to importations from other provinces into Québec. “Unclear” implies no travel history was available and ASR was ambiguous. (C) Travel-related sequences and the TMRCAs of inferred introductions into Québec over time by geographic origin. *Dark densities*: small points indicate sampling dates of sequenced cases with travel history. Large black points indicate the sampling date of the first sequenced case associated with each region. *Pale densities*: small points indicate the TMRCA of the inferred introductions using phylogeny and travel history (thus the date of introduction into Québec will be later). Triangles are the TMRCA of the first estimated introduction from each region into Québec, based on the phylogeny. Asterisks indicate uncertainty due to stem singletons in a large polytomy. The number of introductions is normalized to a relative density within each geographic category (rows).

### Successful cryptic introductions before the first reported case are unlikely

The first confirmed case of COVID-19 in Québec was detected on February 25^th^, but phylogenetic analysis has the potential to infer earlier introduction events. We inferred 13-16 potential introduction events before February 25^th^, based on their time to the most recent common ancestor (TMRCA), of which only 4-5 had reported travel history. The global phylogeny during the first two months of 2020 is undersampled, due to sequencing efforts only beginning to ramp up at that time. This, combined with relatively slow accumulation of mutations by SARS-CoV-2, resulted in many large polytomies (unresolved branchings), making precise inference of introductions challenging. Indeed, ten of these 14-16 early introductions are in polytomies and 53-63% are of unclear origin; thus their true TMRCA is questionable (**Fig. 2C**). The earliest reliably dated introduction has a TMRCA of February 15^th^, from Europe (clustering with sequences from Switzerland), followed by an introduction from the UK with a TRMCA of February 21^st^. We do not reliably detect any introductions arriving in January or early February, which is consistent with a study of samples from patients with flu-like symptoms between November 2019 to early March that did not find any SARS-CoV-2, suggesting that introductions before late February are unlikely (as reported in *Le Devoir*, September 5^th^, 2020; “L’épidémie a Commencé Autour de La Relâche Scolaire,” 2020) and appear not to have given rise to sustained transmission.

### Most introductions occurred after spring break

To test the hypothesis that spring break travel was a major source of viral introductions into Québec, we investigated the Québec transmission lineages with a TMRCA between Feb. 23^rd^ and March 10^th^ and defined them as having been likely introduced during spring break. During this period, there were 70-96 introduction events (only 12-16% of the total), of which 26-36 had recorded travel history (∼37%). This is a conservative estimate since people may have traveled just before or after those precise dates. Also, given the SARS-CoV-2 generation time and incubation period, some introductions during spring break are expected to appear in the counted cases past March 16^th^. The majority of introductions, then, happened after spring break, with 78-82% of TMRCA dates after March 10^th^ (**Fig. 2C**). The USA is a common travel destination for Québecers, where many (known as ‘snowbirds’) have winter homes. The bulk of the USA travel-related cases were detected after the border closed on March 16^th^, and thus were likely part of the repatriation effort. However, the phylogenetically inferred introductions from the USA suggest that these were not as successful as the introductions that happened in early March (the only transmission lineages with >20 viral genomes of US origin arrived before March 15^th^, **Fig. 2C**). The majority of the 41 introductions from other Canadian provinces were not reported in travel history records (38-39 introductions, 93-95%), which is consistent with inter-provincial travel having been common until being discouraged in late March.

### Successful transmission lineages arrived during spring break and spread widely

Of the 555-614 independent inferred introduction events, the majority were unsuccessful: 52-63% were singletons (lineages with only one observed sequence) and 84-93% gave rise to small transmission lineages of less than ten sampled genomes. In contrast, only 5-10 introductions (0.9-1.6% of the total; range of estimates from parsimony and ML) were successful enough to cause more than 50 cases in Québec (**Fig. 3**). The ten introductions inferred by ML events gave rise to 1,414 genomes, or 48% of all sequenced cases (range: 5-10 introductions giving rise to 36-58% of all sequenced genomes in the first wave) until June 1^st^. This overdispersion is more extreme but qualitatively similar to a UK study where the eight largest introductions resulted in >25% of cases (du Plessis et al. 2021). This highly overdispersed transmission lineage size distribution (**Fig. 4A**) is also similar to what was observed in Massachusetts (Lemieux et al. 2021). These results are consistent with an overdispersed reproductive number and superspreading, in which most potential transmission events are unsuccessful but a minority give rise to dozens or hundreds of subsequent infections.

**Fig. 3:**
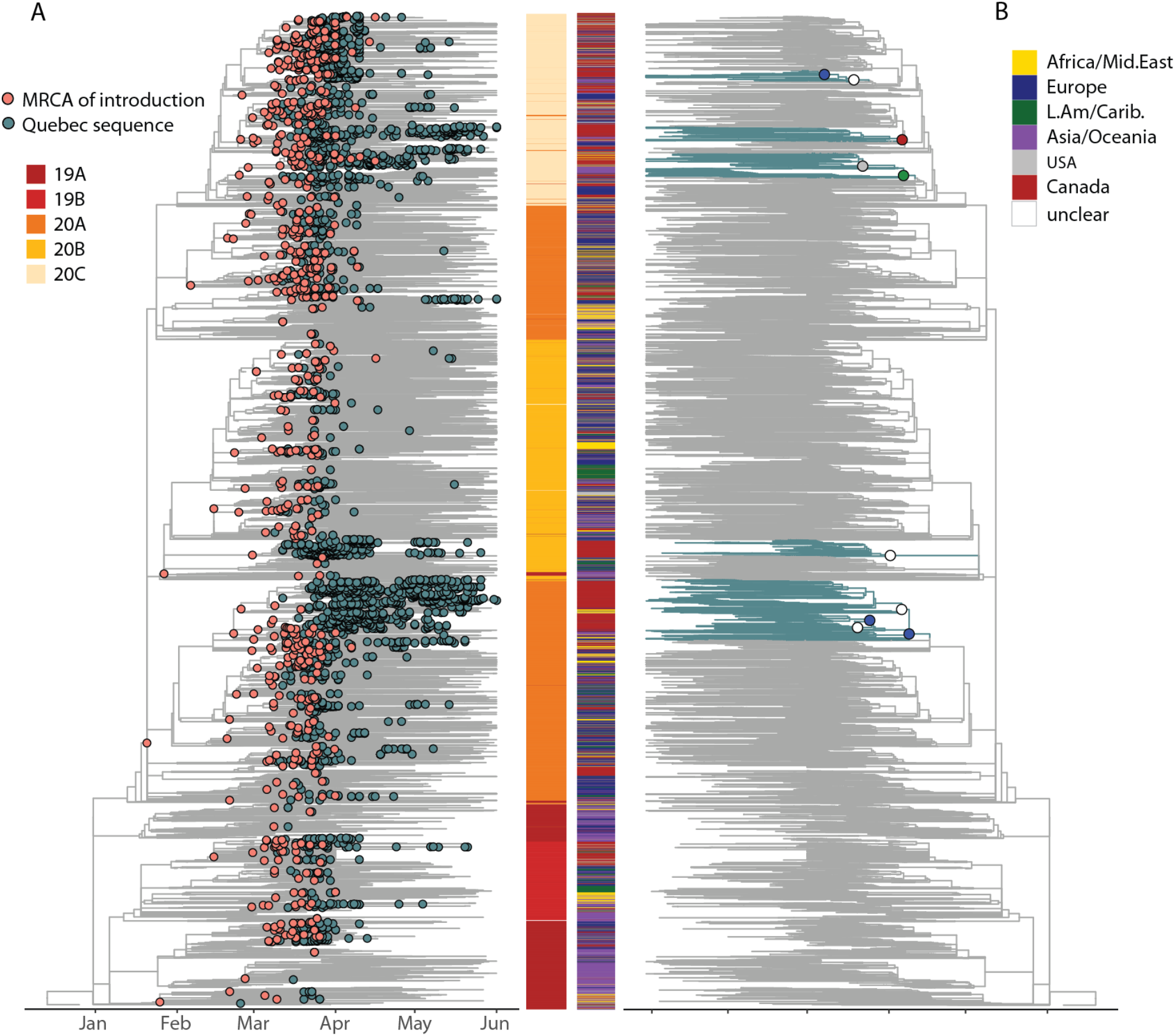
Phylogeny of SARS-CoV-2 genomes sampled from Québec in global context. (A) Pink dots on the time-scaled phylogeny show the most recent common ancestors (MRCA) of introduction events into Québec, inferred with ML ASR. Blue dots are all Québec sequences in the dataset. The heatmap shows Nextstrain clade designations for all sequences in the tree. (B) The same phylogeny, highlighting (blue branches) the ten Québec transmission lineages that gave rise to over 50 sequenced cases. Their introductions (large circles) are colored by their inferred region of origin. The colored heatmap shows the geographic origin of all sequences in the tree.

**Fig. 4.**
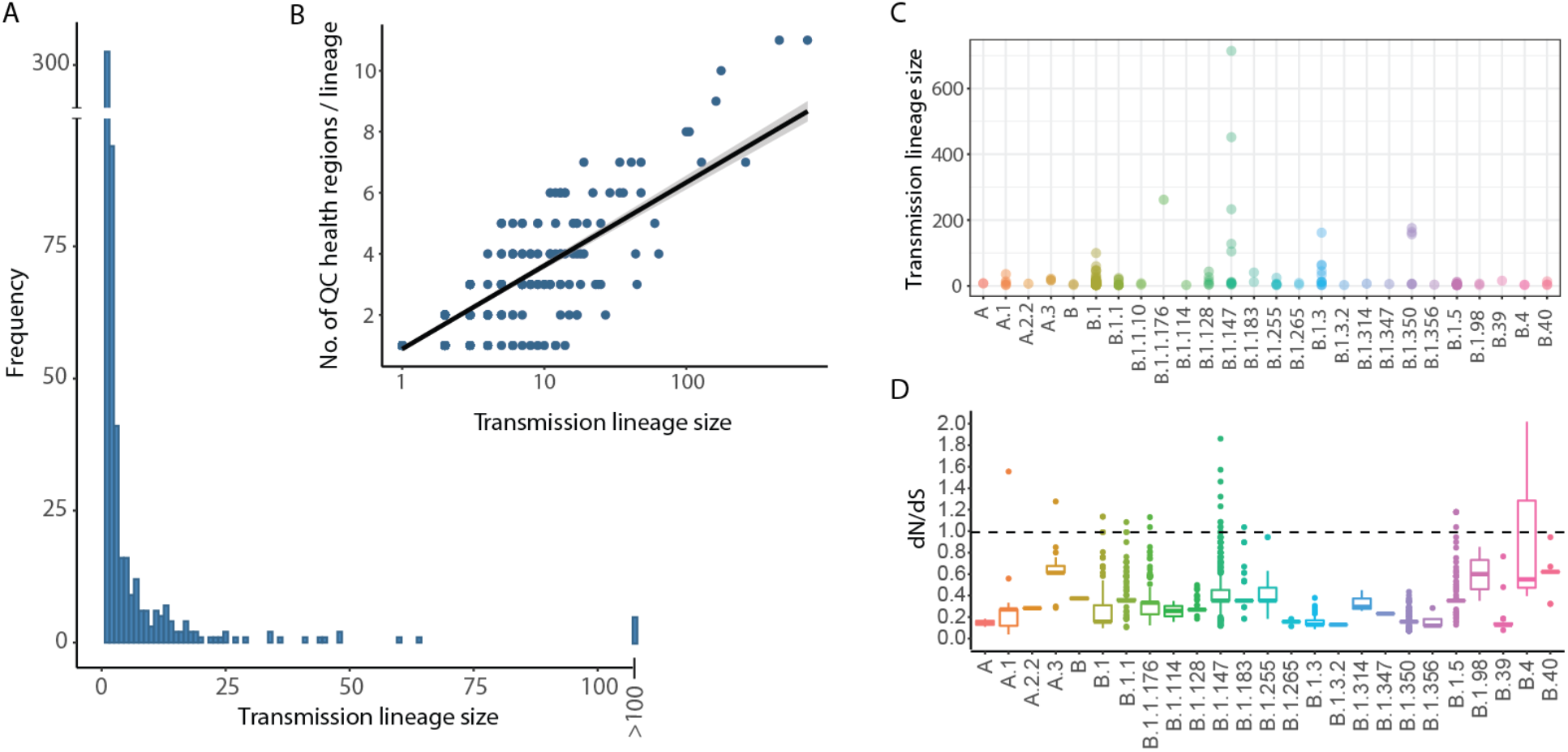
A minority of introduction events are successful and spread across regions. (A) Distribution of Québec transmission lineage sizes, inferred using maximum likelihood ASR, n = 614. (B) Correlation between transmission lineage size and the number of Québec health regions in which it was sampled (Pearson’s r = 0.6, 95% CI = 0.54 −0.64). (C) Transmission lineage sizes generated by each Pangolin viral lineage. Each point represents an independent introduction event into Québec. (D) Estimates of dN/dS for each viral lineage in Québec. For each viral lineage, the boxplots represent the distribution of dN/dS across sampled genomes compared to the ancestral reference genome (Wuhan-1; Genbank Accession MN908947.3).

Larger transmission lineages tended to be sampled across more regions in Québec (**Fig. 4B**), indicating that the success of these lineages was not due to local outbreaks but rather to wide geographic spread across the province. The viral lineages that spread the most throughout Québec (*i*.*e*. being found in more than ten health regions) were B.1, B.1.5, and B.1.147 (**Fig. S2**). Viral lineage B.1.147 stayed mostly in the more populous southern regions of the province while genomes of B.1 were found in almost all regions (**Fig. S2** and **Fig. S3**). Viral lineage B.1.5 first arrived after spring break and was introduced 41 times across Québec but was not successful at generating transmission lineages of over 12 sampled genomes, and was no longer observed by June 1^st^ (**Fig. S1** and **Fig. S2**). In contrast, B.1.147 was introduced half as many times (19 introductions) but these events tended to have occurred earlier in spring break (**Fig. S1**).

The most successful Québec transmission lineages were caused by B.1.147, B.1.350, B.1.3, and B.1, each of which was introduced multiple times (**Fig. 4C, Fig. S3**, and **Fig. S4**). These viral lineages had evolutionary rates comparable to other lineages in Québec (∼6×10^−4^ substitutions per site per year; **Fig. S5**), somewhat slower but in the range estimated from other studies (Duchene et al. 2020). To quantify variation in the strength of natural selection on these lineages, we calculated the nonsynonymous to synonymous substitution ratio (dN/dS) between all pairs of genomes within a viral lineage (**Fig. 4D**). There was a modest positive correlation between a viral lineage’s dN/dS and its average transmission lineage size (Pearson’s *R*^*2*^ = 0.12, permutation test *P* = 0.0002). Much of this correlation is driven by lineage B.1.147 (**Fig. 4C,D**) and could be explained by the accumulation of low-frequency, slightly deleterious nonsynonymous mutations at the tips of a rapidly expanding clade (Rocha et al. 2006; Charlesworth and Eyre-Walker 2008), which is also consistent with strongly negative Tajima’s *D* values (**Fig. 1C**). Together, these results suggest rapid population growth of the most successful lineages.

The ten largest transmission lineages likely arrived during spring break (all TMRCA 95% HPD intervals overlap with spring break) and were still detectable in late May (**Fig. 5**). The median effective reproductive numbers (*R*_*e*_, estimated by phylodynamic analysis) for the two largest transmission lineages were estimated in the range of 2-3, consistent with exponential growth (**Fig. 5**). The two transmission lineages of ∼60 cases caused by introductions of B.1 and B.1.3 had higher *R*_*e*_ values, potentially due to their rapid spread in long-term care facilities. The B.1 transmission lineage that spread mostly in a care facility in the city of Laval (McKenzie 2020), is a particularly striking example, where the median age jumped to 83 years old (IQR: 71 to 89 years) after a likely introduction by a person in their forties (**Fig. S6**). This outbreak was brought under control, and then no sequences were detected past early May (**Fig. 5; Fig. S6**). The self-isolation mandates for arriving travelers (Québec’s orders on March 11^th^ and federal mandatory quarantine orders on March 25^th^) appear to have been effective, such that the mean transmission lineage size before March 11^th^ was 22 cases (+/-85 SD) and only 3 cases (+/-5.1 SD) after March 11^th^. After the federal quarantine orders, 70% of introductions were singletons and only four gave rise to 10 or more sampled genomes. The TMRCA of the last introduction event was April 16^th^, 2020.

**Fig. 5.**
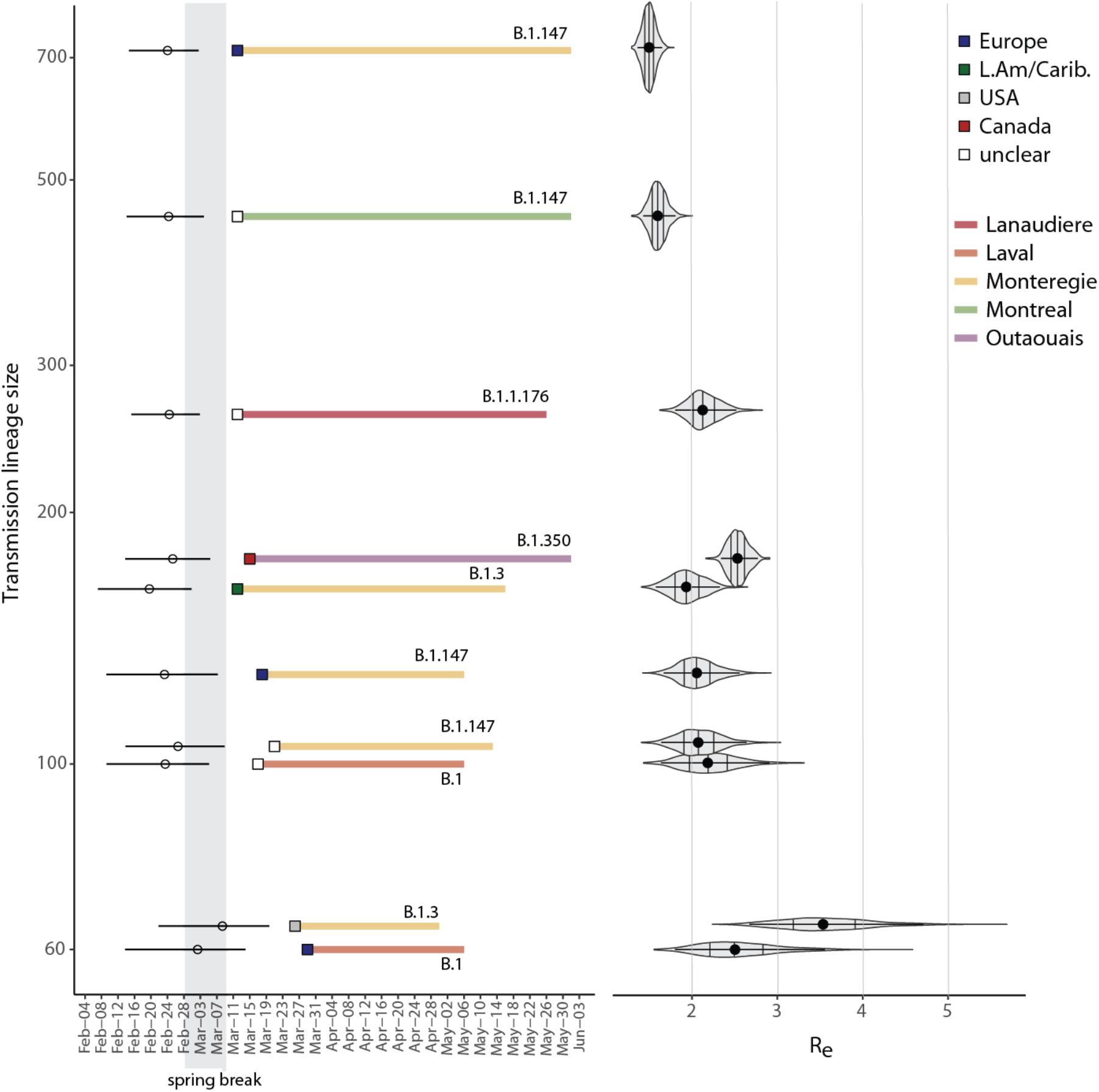
Introduction and duration of the ten largest Québec transmission lineages each responsible for 50 or more cases. Timelines are coloured by the health region in which each Québec transmission lineage predominantly spread, and which SARS-CoV-2 lineage (Pangolin nomenclature) was responsible. Squares represent the inferred origin of the introduction event, open circles are median and HPD 95% interval of TMRCA of each lineage. Right: median and HPD 95% interval of the effective reproductive number, R_e_, for each lineage, estimated using BEAST (Methods). These transmission lineages were inferred by ML ASR.

### Mutation and founder effects on transmission lineage size

Finally, we considered the extent to which the success of an introduction event could be explained by founder effects and adaptive mutations. To investigate the role of specific mutations, we defined nine lineage-specific single nucleotide variants (alleles) present in all members of each viral lineage, and tested their associations with transmission lineage size. We found that mutation D614G in the Spike protein (genome position A23403G) was present in all ten of the most successful introduction events into Québec (**Fig. S7**) and generally dominated our sampled sequences (**Fig. 6A**). Independent introductions of viral lineages with the derived G allele gave rise to a mean transmission lineage size of 6.6 cases, compared to 3.4 for the ancestral D allele; however, this difference is not statistically significant (**Fig. S7**). In contrast, derived nonsynonymous mutations in three consecutive nucleotide sites (28881-3) spanning two codons in the nucleocapsid (N) protein were significantly associated with smaller transmission lineage size (**Fig. S7**) and were less represented in our sequences (**Fig. 6A**).

**Fig. 6.**
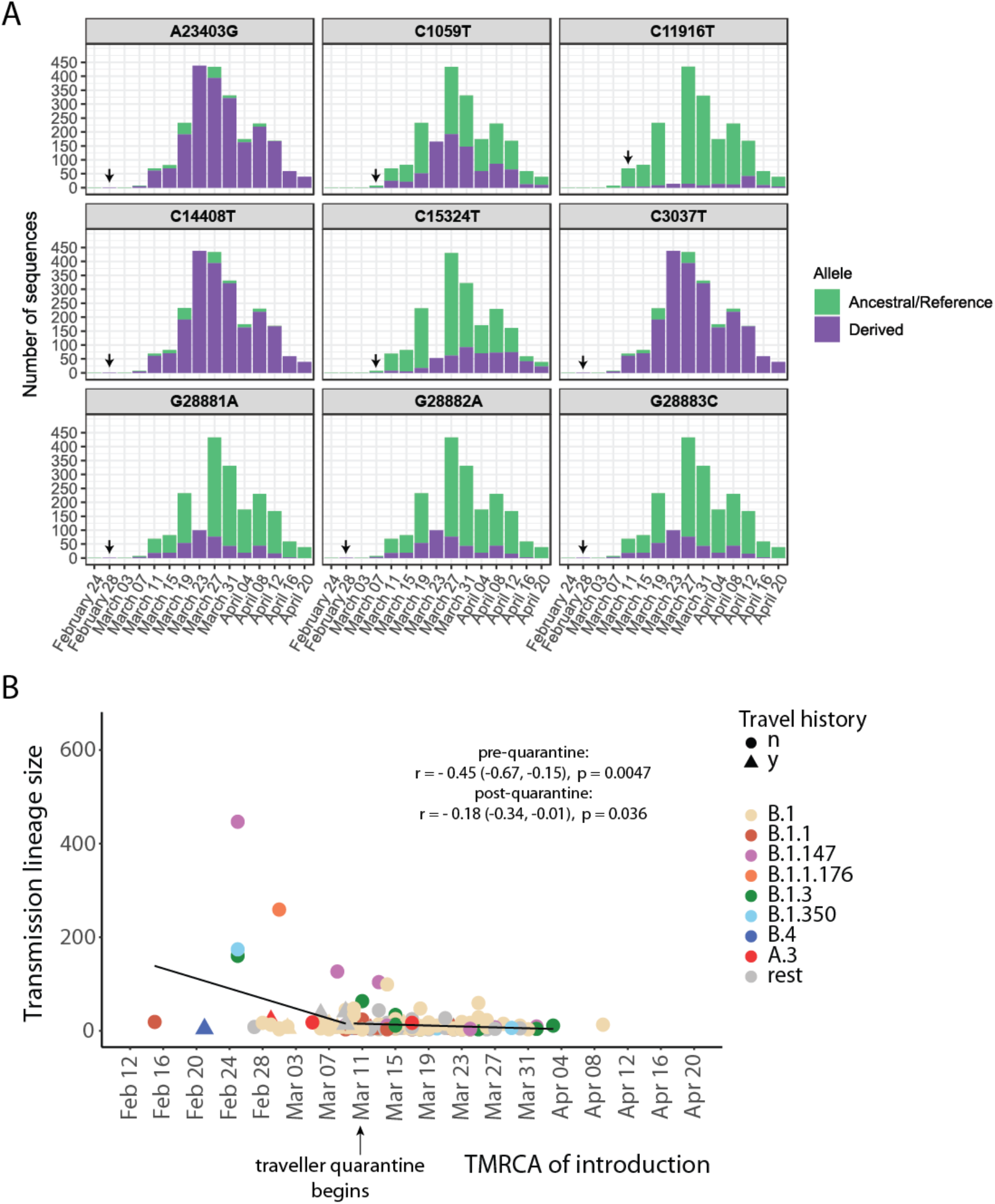
Arrival of SARS-CoV2 lineages and transmission-associated mutations over time. (A) Number of consensus sequences including each lineage-defining mutation, named by alternative nucleotides and genome position. The first detected occurence of each derived allele is indicated by an arrow. (B) Introductions that generated transmission lineages of >2 genomes as a function of the TMRCA of the lineage (inferred by ML ASR), colored by viral lineage name and annotated if the introduction had reported travel history (y) or not (n). Pearson correlation coefficients, 95% CIs and p-values are shown for pre-and post-quarantine time periods. Overall: *r* = −0.33 (−0.46, −0.19), *p* < 0.0001, no line shown. Note that the last inferred introduction event had a TMRCA of April 16^th^, not shown here because it was a singleton.

If founder effects also played a role in determining successful transmission, we would expect the earliest introduction events to give rise to larger transmission lineages. Consistent with founder effects, we observed a significant negative correlation between inferred arrival time and transmission lineage size. This negative correlation was strongest before quarantine measures began on March 11th, suggesting that that founder effects occurred independently and could be decoupled from the effects of quarantine measures (**Fig. 6B**). The negative correlation was detectable but weak after March 11th, suggesting a strong effect of quarantine measures. We also note that most of the early, successful introduction events had no reported travel history, highlighting the importance of phylogenetic analysis in identifying them. Therefore, while we cannot exclude a role of specific mutations affecting transmission, lineage success in Québec’s first pandemic wave can most parsimoniously be explained by a combination of founder effects and effective public health measures.

## Conclusions

Québec is unique among Canadian provinces for its early spring break, which resulted in many returning travelers before border closures and quarantine measures were enacted. Our results are consistent with mathematical modelling results (Godin et al. 2021) suggesting that Québec’s large and severe SARS-CoV-2 epidemic was driven by this early spring break. Even if most introduction events likely occurred well after spring break, those that arrived during or just after spring break generated the largest transmission lineages. Before quarantine and other public health measures were in place, ten introductions that arrived during spring break gave rise to hundreds of subsequent infections and spread widely across Québec. While hundreds of introduction events continued to occur after spring break, these spread much less widely, likely due to effective public health measures. This scenario closely mirrors the early SARS-CoV-2 introductions into the UK, which also spread widely and proved hard to eliminate (du Plessis et al. 2021).

Our phylogenetic analysis is generally concordant with self-reported travel history, but also revealed a large number of introduced European SARS-CoV-2 viral lineages that were not apparent from travel history. Québec is, thus, similar to other East Coast North American epidemics, such as Boston (Lemieux et al. 2021) and New York (Gonzalez-Reiche et al. 2020), that were primarily seeded by European lineages, in contrast to more Asian lineages on the West Coast (Worobey et al. 2020). The Québec sequences broadly clustered across the global phylogenetic tree, representing most of the known diversity, with an under-representation of early-branching Asian lineages. Like other phylogenetic studies, ours is limited by sampling: we cannot reliably detect introduction events from countries poorly represented in public databases, nor have we sequenced all SARS-CoV-2 infections in Québec. We were able to sequence ∼6% of positive cases, putting our effort nearly on par with other leading genomic surveillance projects (*e*.*g*. currently ∼8% in the UK; https://www.cogconsortium.uk/). Nevertheless, our estimate of at least 500 independent introduction events is almost certainly an underestimate. This highlights the need for sustained genomic surveillance efforts.

Although it is notoriously difficult to disentangle demographic factors from fitness effects of viral mutations (Grubaugh, Hanage, and Rasmussen 2020), our results are consistent with a mild (not statistically significant) transmission advantage of the D614G Spike mutation, as observed elsewhere (Volz et al. 2021). We also identified three adjacent derived mutations in the N protein associated with smaller transmission lineage size. These mutations (nucleotide positions 28881-28883) have been reported before, but their functional significance remains unclear (Garvin et al. 2020) and could warrant further study. While these mutations may have played some role in affecting transmission in Québec, the differential success of introduced lineages is parsimoniously explained by founder effects, such that the first lineages that arrived tended to be successful. The recent success of lineage B.1.1.7, which spread in the UK and elsewhere despite competition from previously established lineages, cannot be easily explained by founder effects (Davies et al. 2021). Nevertheless, founder effects and other demographic factors must be carefully considered when inferring a transmission advantage of viral lineage of interest.

Consistent with superspreading dynamics, we observed an overdispersed distribution of introduced transmission lineage size: most introduction events went extinct, while only 5-10 introductions (<2%) gave rise to at least one third of sequenced cases. Viral lineages that were introduced after the self-isolation mandate for travellers were largely unsuccessful at generating large transmission lineages, despite repeated introductions into Québec. Although our province-wide sampling was not designed to focus on specific outbreaks, they are reflected in our dataset. For example, one introduction of viral lineage B.1 during spring break quickly spread from younger to older individuals in a long-term care facility. This example mirrors the broader trajectory of the Québec epidemic in the first pandemic wave. Our study demonstrates the importance of timely public health actions during the early phases of a pandemic and how they shape the dynamics, size, and geographical spread of a novel pathogen.

## Data Availability

All reported genome sequences have been deposited in GISAID under accession numbers listed in Table S1.

https://www.gisaid.org/

## Acknowledgements

We thank all the authors, developers, and contributors to the GISAID database for making their SARS-Cov-2 sequences publicly available. We are grateful to the molecular biology team of the public health laboratory of Québec (LSPQ) including Lyne Desautels, Martine Morin and Mélanie Côté for thoroughly collecting and aliquoting all the COVID-19 positive samples. We would like to thank Marie-Michelle Simon and Patrick Willet for technical assistance on sample processing and Alexandre Belisle for automation assistance at the McGill Genome Center. Illumina and MGI sequencing was performed by Janick St-Cyr and Pierre Lepage. We thank members of the public health surveillance committee for SARS-CoV-2 for their contribution to the validation of data and their review of the manuscript and the team *Immunisation et infection nosocomiale* from the Public Health Institute of Québec. The work was supported by the McGill Genome Center and the Canadian Center for Computational Genomics, two Genomics Technology Platforms (GTPs) supported by the Canadian Government through Genome Canada and a CFI grant 33408 to JR and GB. This study was also funded by the Sentinelle COVID Québec variant network led by the Laboratoire de Santé Publique du Québec (LSPQ) in collaboration with Fonds de la Recherche du Québec-Santé (FRQS) and Genome Québec, and supported by the Ministère de la Santé et des Services Sociaux (MSSS), the Ministère de l’Économie et Innovation (MEI) and Genome Canada to SM and MR under the umbrella of the Canadian COVID Genomic Network (CanCOGeN). Data analyses were enabled by compute and storage resources provided by Compute Canada and Calcul Québec.

## Methods

### Sampling and sequencing

COVID-19 positives cases were selected from all nasopharyngeal swabs sent to the Public Health Laboratory of Québec (Laboratoire de Santé Public du Québec, LSPQ) from the beginning of the pandemic until June 1^st^, 2020. To protect patient confidentiality, in the publicly released data the first accurate date of sampling is set to March 10th, 2020. All samples taken before that are set to March 1^st^, and their real sampling dates are between February 25th and March 9th, 2020. The true sampling dates were used in phylogenetic analyses described below.

Total nucleic acid extraction was performed with the NucliSENS EASYMAG automated platform on 200 μL of nasopharyngeal swabs. The presence of SARS-CoV-2 was assessed by a qPCR diagnostic test targeting genes E and N (LeBlanc et al. 2020). Targeted SARS-CoV-2 amplification, library preparation, and sequencing were performed at the McGill Genome Centre as follows. Briefly, RNA samples were processed in a 96-well plate format, including positive and negative controls on each plate. A targeted amplification strategy was used based on the ARTIC V3 primer scheme (https://github.com/artic-network/artic-ncov2019) using the V3 primers only without adding the redundant V1 primers. For primer pairs 5, 17, 23, 26, 66, 70, 74, 91, 97, and 64, for which a lower coverage was observed, a separate additional low amplification (LA1) pool was prepared to increase the number of reads in the corresponding region. For post-PCR cleanup, pools 1 and 2 were combined, while pool LA1 was cleaned up separately, quantified, and added to the combined pools 1+2 in equimolar concentration. Samples from plates 1-4 were prepared for Nanopore sequencing as described : https://www.protocols.io/view/sars-cov-2-mcgill-nanopore-sequencing-protocol-sup-bjajkicn. For Nanopore sequencing, we used native barcodes on pooled amplicons and loaded 20-40 ng of library onto the flow cell. Samples from plates 5-8 were prepared for Nextera Flex Illumina sequencing as described: https://www.protocols.io/view/sars-cov-2-mcgill-nextera-flex-sequencing-protocol-bisbkean. For samples sequenced on Illumina, the Nextera Flex kit was used starting from 150 ng of DNA following the procedure from the manufacturer. Plate 9 was sequenced using both Nanopore and Illumina technologies, as well as by applying the Cleanplex assay by Paragon Genomics, followed by MGI sequencing. For samples that were sequenced more than once, the data with the higher coverage was used to generate consensus sequences and subsequent phylogenetic analysis. For the Cleanplex assay, we used the Cleanplex for MGI SARS-CoV-2 research panel by Paragon Genomics. The assay utilizes 343 primer pairs tiled across the viral genome as described (Li et al. 2020). The manufacturer’s protocol (UG4002-1) was used, with the modification of increasing the multiplex PCR cycle number to 16 in order to improve the sequencing of samples with qPCR cycle threshold (Ct) values of >29, followed by sequencing on an MGI DNBSEQ G400 instrument.

### Basecalling and consensus sequence generation

All samples were aligned to the reference genome of the Severe Acute Respiratory Syndrome Coronavirus-2 isolate Wuhan-Hu-1 (GenBank Accession MN908947.3). Aligned reads were then used to produce a consensus sequence using pipelines based on the Artic Network novel <u>coronavirus bioinformatics protocol</u>. A brief description of the pipeline, including software packages and important parameters, is provided for each sequencing platform below.

Datasets produced using the Nextera Flex Illumina protocol were first filtered to remove any host reads. To do so, reads were aligned to a hybrid reference including SARS-CoV-2 (MN908947.3) and GRCh38 using bwa-mem (v0.7.17). Any reads mapping to a region of the human reference with a mapping quality of zero or more were removed from the dataset. After filtering out host reads, the remaining reads were trimmed using cutadapt (v2.10), then aligned to the SARS-CoV-2 reference (MN908947.3) using bwa-mem (v0.7.17). After alignment, reads were filtered using sambamba (v0.7.0) to remove paired reads with an insert size outside of the 60-300bp range, as well as any unmapped reads, secondary alignments and reads that did not match the FR/RF orientation. iVar (v1.3) was used to trim any remaining primers. Samtools (v1.9) was used to produce a pileup which was then used as input by iVar (v1.3) to create a consensus sequence for regions with a minimum of 10x depth, using reads with a Q score >20 and a minimum allele frequency of 0.75. A full description of the process can be found here: https://c3g.github.io/covseq_McGill/SARS_CoV2_Sequencing/Illumina_overview.html

Datasets produced using the CleanPlex MGI protocol were processed using the same pipeline as Illumina Nextera Flex samples, except that Artic Network primers and amplicon data was changed to the corresponding CleanPlex information. A full description of the process can be found here: https://c3g.github.io/covseq_McGill/SARS_CoV2_Sequencing/MGI_overview.html

Raw data produced using Nanopore sequencing was basecalled using guppy (v3.4.4) with a High-Accuracy Model (dna_r9.4.1_450bps_hac). Reads were de-multiplexed using guppy barcodes (v3.4.4), requiring barcodes on both ends. Reads were filtered by size to remove anything outside of the 400-700bp range using the ARTIC Network ‘guppyplex’ tool. Reads were aligned with minimap2 (v2.17), then filtered to remove incorrect primer pairs and randomly downsample high-depth regions to keep 800x depth per strand using the ARTIC network framework. Nanopolish (v0.13.1) was used to call variants in regions with a minimum depth of 16x and a flank of 10bp. After masking regions with coverage below 20x, the called variants were used to generate a consensus sequence using bcftools (v1.9) consensus. A full description of the process can be found here: https://c3g.github.io/covseq_McGill/SARS_CoV2_Sequencing/ONT_overview.html

For samples sequenced with two or more technologies, all datasets were processed separately using the methods described above. The resulting consensus sequences were compared to keep only the most complete consensus for downstream analyses, as determined based on the number of missing bases (Ns). The consensus sequences were deposited in GISAID under accession number listed in **Table S1**.

### Phylogenetic analysis

Raw and time-scaled phylogenomic trees were built using the NextStrain pipeline (https://github.com/nextstrain, version 1.16.2) installed in a conda environment (https://github.com/conda/conda, version 4.8.3) (Hadfield et al. 2018). This pipeline uses the Augur toolkit (https://github.com/nextstrain/augur, version 7.0.2) to filter, align/mask genomic sequences, build trees (divergence and time-scaled) and produce an output file processed by the Auspice web interface (https://github.com/nextstrain/auspice, version 2.16.0) to explore phylodynamic and phylogenomic data. Augur removed all sequences shorter than 27,500 bp and sampled after June 1^st^, 2020. The Augur/align module was then called to execute the multiple sequence alignment with MAFFT (https://github.com/GSLBiotech/mafft, version v7.463) using Wuhan-1 (Genbank accession MN908947) as a reference genome. The final alignment was masked at the beginning (first 100 sites) and end (last 50 sites), and at positions 18529, 29849, 29851 and 29853 (sites of known low sequencing quality and homoplasies). We then used Augur to select sequences from GISAID that were most similar to our 2,921 Québec sequences. These global context sequences were then grouped by country/month in order to keep a maximum of 100 sequences and 5 identical sequences per country-month combination.

We used IQ-TREE (http://www.iqtree.org/, version 1.6.12) to construct a phylogenetic tree of Québec only sequences and another tree of Québec and global context sequences, with the GTR substitution model. Branch lengths, sampling dates and ancestral states (geographic regions, nucleotides and amino acids sequences) at internal nodes were inferred with the Augur/refine and Augur/traits modules by calling TreeTime (https://github.com/neherlab/treetime, version 0.7.5) (using the same default parameters as those chosen in public builds; https://github.com/nextstrain/ncov) (Sagulenko, Puller, and Neher 2018). Finally, the Augur/export module exports a single compiled results file required for data visualization in Auspice. All Nexstrain analyses were executed on a 64-bit CentOS server version 7.4.1708 using 40 CPUs.

Clade assignment was done during the Nextrain build. As input, the Augur/clades module uses the phylogenetic tree, the observed and inferred nucleotide sequences at each node and a clade configuration file. In this clade file, every single clade value is associated with a specific combination of position/nucleotide variant. As an alternative clade assignment scheme, we also used the Phylogenetic Assignment of Named Global Outbreak LINeages (Rambaut et al. 2020) combined with lineages (https://github.com/cov-lineages/lineages) version 2020-05-09 (Pangolin 2.3.2 and pangoLearn 2021-02-21; <u>cov-lineages.org</u>).

To infer introduction events into Québec (QC), we used discrete character ancestral state reconstruction (ASR) to infer non-QC and QC nodes in the global context time tree. Three methods were implemented in *R* using either: (1) maximum likelihood (*ace* function from *ape* package v5.4-1, assuming the equal rates model), or unordered Fitch parsimony implementing either with (2) delayed (DELTRAN) or (3) accelerated (ACCTRAN) transformation algorithms in order to deal with ambiguous nodes (*fitch*.*mvsl* function from package *mvSLOUCH* v2.6.1). With the reconstruction, we assigned nodes to the QC state when supported by ≥50% (with ML) or 1 (with parsimony) of the state assignment. By starting at the root of the tree we identified transition nodes, i.e. as non-QC nodes that immediately precede a QC node or leaf, by testing all non-QC nodes for any QC children and whether or not they (or their parent node) have already been identified as a transition node. Transition nodes that were also polytomies were flagged. Note that these methods likely underestimate the number of introductions. The non-QC to QC transitions were collected and their most basal QC leaf (or leaves) are recorded. These candidates were then cross-checked with travel history data and were only recorded if at least one had travel-history. In the case of a polytomy with multiple basal QC sequences, only one was chosen by the shortest branch length. If no travel history was available then the closest outgroup of the introduction event was used to assign the likely origin of the introduction event. Non-QC to QC transition nodes were labeled as likely introduction events, and the descendants of these identified transition nodes were used to define QC transmission lineages. The date of the non-QC to QC transition node was used as the TMRCA of the introduction event. For the largest QC transmission lineages (containing >50 cases) the TMRCA was also inferred using BEAST (see below). Phylogenetic visualizations and dataset manipulation were done in R using a suite of packages: *ape, phylotools, phytools, phangron, tidyverse* and *ggtree*.

### Phylodynamics

The molecular clock signal was assessed by plotting the root-to-tip phylogenetic distance against time using TempEst (Rambaut et al. 2016). The largest (>50 cases) QC transmission lineages were analyzed using Bayesian phylogenetic tree reconstruction with Markov chain Monte Carlo (MCMC) implemented in BEAST v2.6.2 (Bouckaert et al. 2019) with the Birth-Death Skyline (BDSKY) model, assuming a gamma distributed Hasegawa-Kishino-Yano (HYK) nucleotide substitution model (with a uniform distribution 0.25 [0,1] of the nucleotide frequencies, a lognormal 2 [0, ∞] for *ϰ*, and a *γ* count of 4 with an exponential distribution 0.5 [-∞, ∞]) and a strict molecular clock (0.8 x 10^−3^ substitutions/site/year). Using this model, we estimated the effective reproduction number (*R*_*e*_), TMRCA, and sampling proportion. The prior for the reproduction number was a lognormal distribution (initial = 2 [0,10], M=0, S=0.5), origin was a normal distribution (mean=0.1, σ=0.05, initial 10[0,∞]), the rate of becoming infectious was a normal distribution (mean=10, σ=1.3, initial =1[0,∞]), and sampling rate was a beta distribution (*α*=1, *β*=5, initial= 0.01[0,1]). All MCMC analyses were run with 50 million generations and sampling every 50,000 steps for lineages with >100 cases and 30 million generations and 30,000 steps for lineages < 100 cases. Convergence was achieved when all posteriors gave effective sample sizes (ESS) > 300 with 10% burn-in.

### Calculation of population genetic metrics

We calculated Tajima’s *D* to infer changes of the viral effective population size and deviation from a standard neutral evolutionary model. We separated the data into eight time periods of two weeks between February 20, 2020 and June 10, 2020. For each time period, we randomly sampled 20 viral consensus sequences to calculate Tajima’s *D*, and repeated this procedure 99 times to obtain confidence intervals. We calculated both a combined value of *D* across all sequences, and a separate estimate for each Pangolin viral lineage. Lineages or time bins with fewer than 20 sequences were discarded. We calculated *D* as described (Tajima 1989):

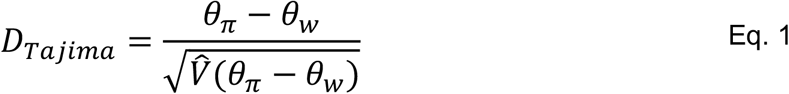

where the 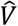 denotes the expected sampling variance of (θ_π_ – θ_*w*_). θ_π_ is the nucleotide diversity, calculated based on the average number of pairwise differences among consensus sequences:

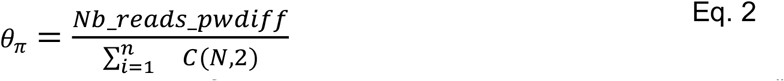

where *n* is the genome length, *N* is the number of consensus sequences, *C*(*N*, 2) is the choose() function which calculates the number of pairs of consensus sequences in a set of size *N* and *Nb_reads_pwdiff* is the number of pairwise nucleotide differences. Because pairwise differences are maximized when there are intermediate-frequency mutations, *θ*_*π*_ is more sensitive to intermediate-frequency mutations. *θ*_*w*_ is another estimator of the nucleotide diversity which is calculated based on the number segregating sites and is sensitive to low-frequency mutations:

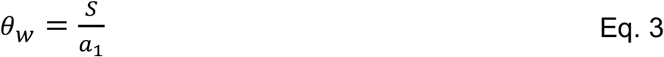

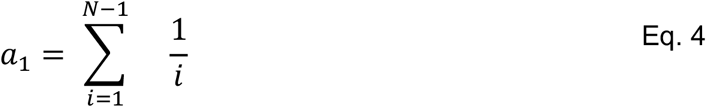

where *S* in the number of segregating sites, *a*_1_is a normalizing factor for the sample size of consensus sequences (*N*).

We also estimated *dN/dS*, the ratio of non-synonymous (*dN*) and synonymous substitutions rates (*dS*), by comparing consensus sequences to the reference genome (Genbank MN908947.3) allowing us to infer changes in selective pressures at the protein level.

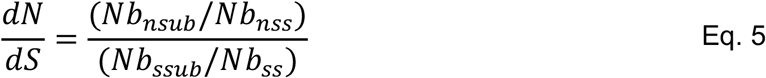

where *Nb*_*nsub*_ is the number of non-synonymous substitutions, *Nb*_*nss*_ is the number of nonsynonymous sites, *Nb*_*ssub*_ is the number of synonymous substitutions, and *Nb*_*ss*_ is the number of synonymous sites. We only considered consensus sequences with more than 1 synonymous mutation to be able to attribute finite values to *dN*/*dS*. These analyses were coded in R.

## Supplementary Figures

**Fig. S1.**
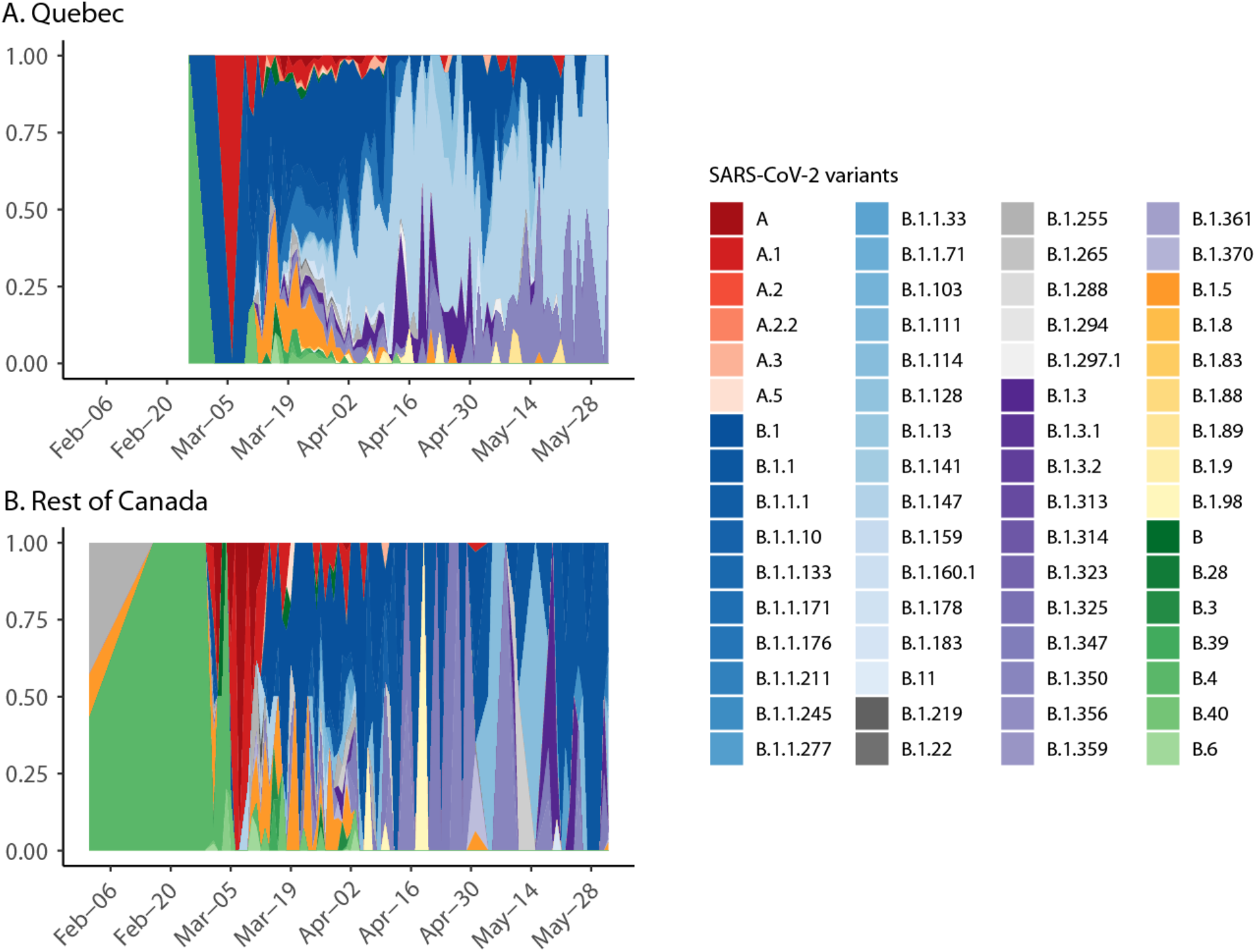
SARS-CoV-2 clades observed over time in (A) Québec and (B) the rest of Canada. Frequencies of named SARS-CoV-2 variants (Pangolin nomenclature; cov-lineages.com) of sampled Québec genomes over time.

**Fig. S2.**
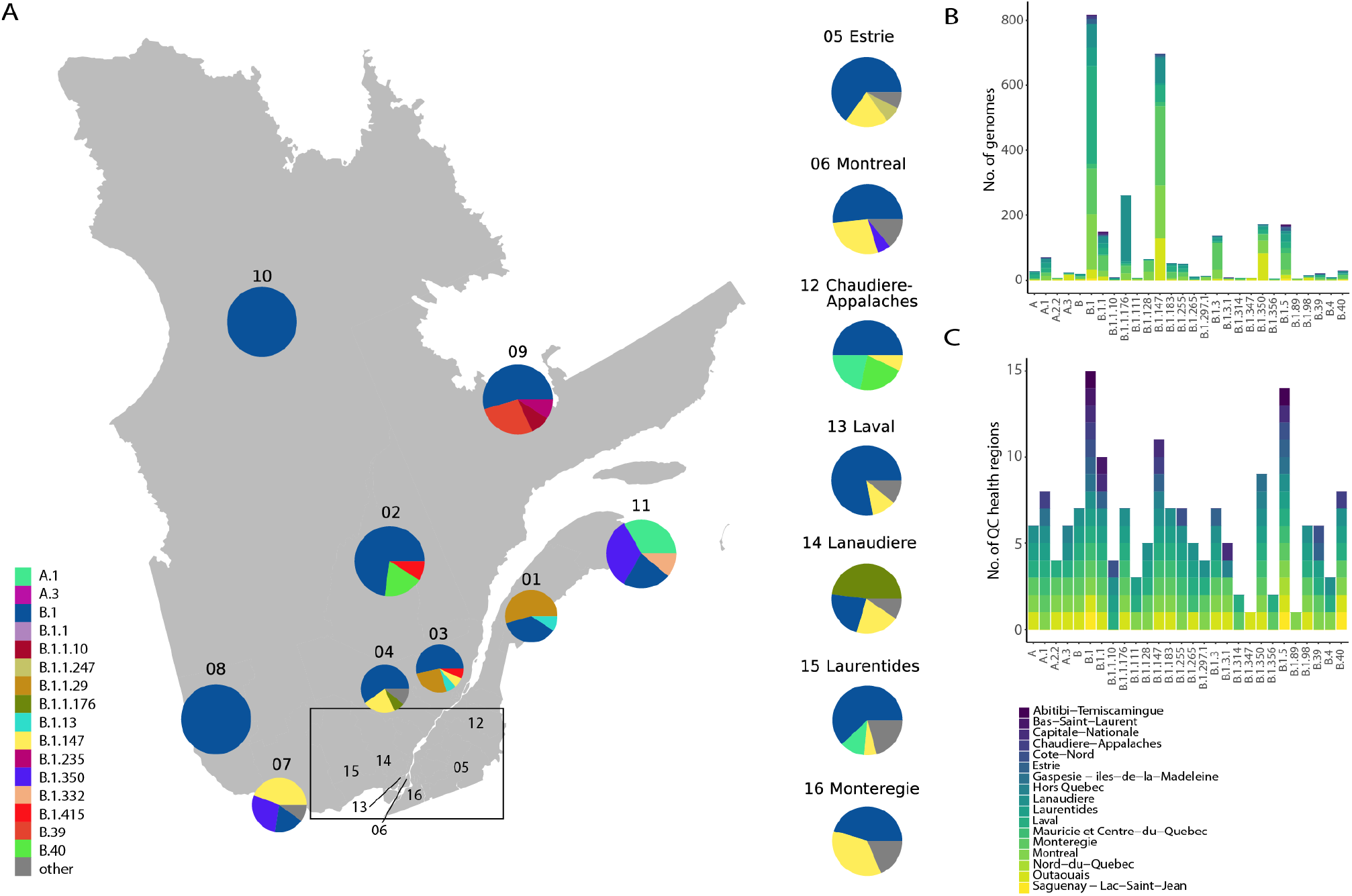
Geographical spread of viral lineages in Québec. (A) Regional distribution of viral lineages, up until June 1st, 2020. (B) Frequency of viral lineages that have >3 genomes in the Québec dataset per health region. (C) Number of Québec health regions in which each named viral variant (Pangolin nomenclature) was found. Québec health regions not named in figure: 01-Bas-Saint-Laurent, 02-Saguenay-Lac-Saint-Jean, 03-Capitale-Nationale, 04-Mauricie-et-Centre-du-Québec, 07-Outaouais, 08-Abitibi-Témiscamingue, 09-Côte-Nord, 10-Nord-du-Québec, 11-Gaspésie-Iles-de-la-Madeleine. Note: Hors-Québec represents non-residents (visitors) who tested positive.

**Fig. S3.**
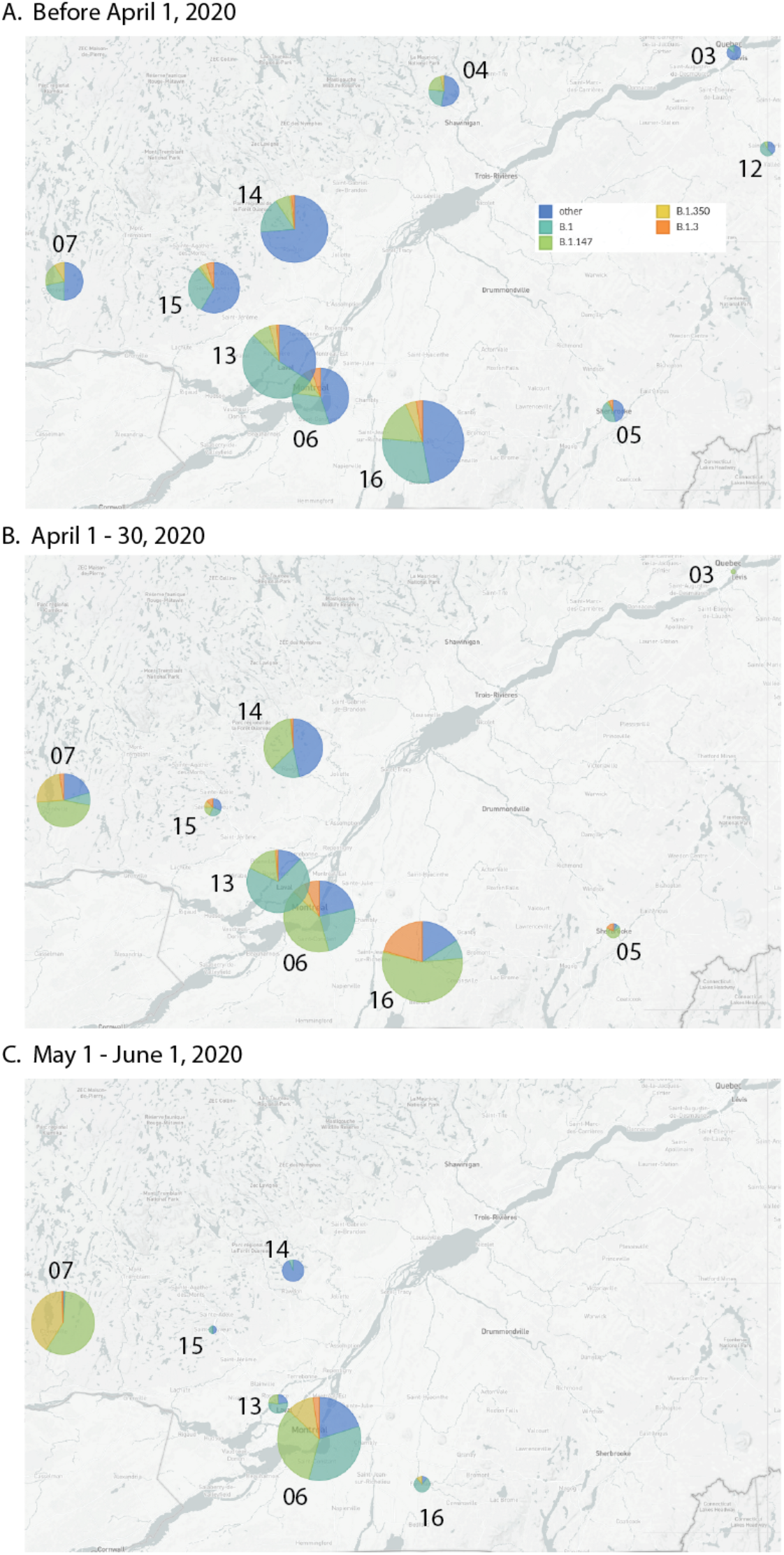
Temporal variation of geographical spread of the most successful viral lineages in the most populous Southern regions of Québec. Maps with pie charts showing the frequencies of viral lineages (A) up to April 1st, during exponential increase of the epidemic, (B) during the plateau of epidemic, and (C) during the decline period. Pie charts are sized proportionally to the total number of sequences sampled in the health region. The Québec health regions depicted here are: 03-Capitale-Nationale, 04-Mauricie-et-Centre-du-Québec, 05-Estrie, 06-Montréal, 07-Outaouais, 12-Chaudière-Appalaches, 13-Laval, 14-Lanaudière 15-Laurentides, 16-Montérégie

**Fig. S4.**
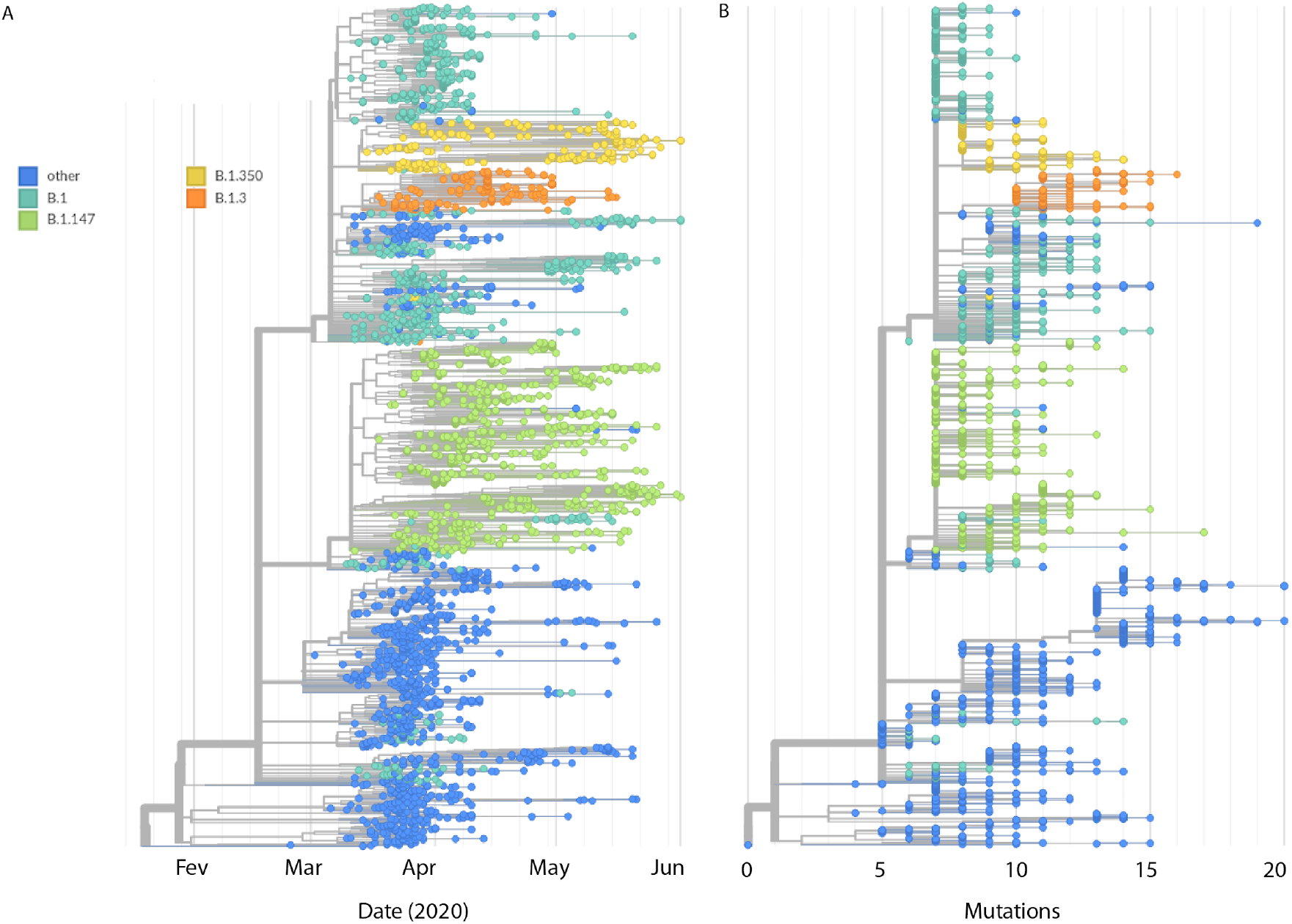
Phylogeny of SARS-CoV-2 genomes sampled in Québec, up to June 1st, 2020. (A) Maximum-likelihood time-scaled phylogenetic tree of all 2,921 Québec sequences from this study. The most successful viral lineages are annotated by color. (B) Divergence tree of the same dataset.

**Fig. S5.**
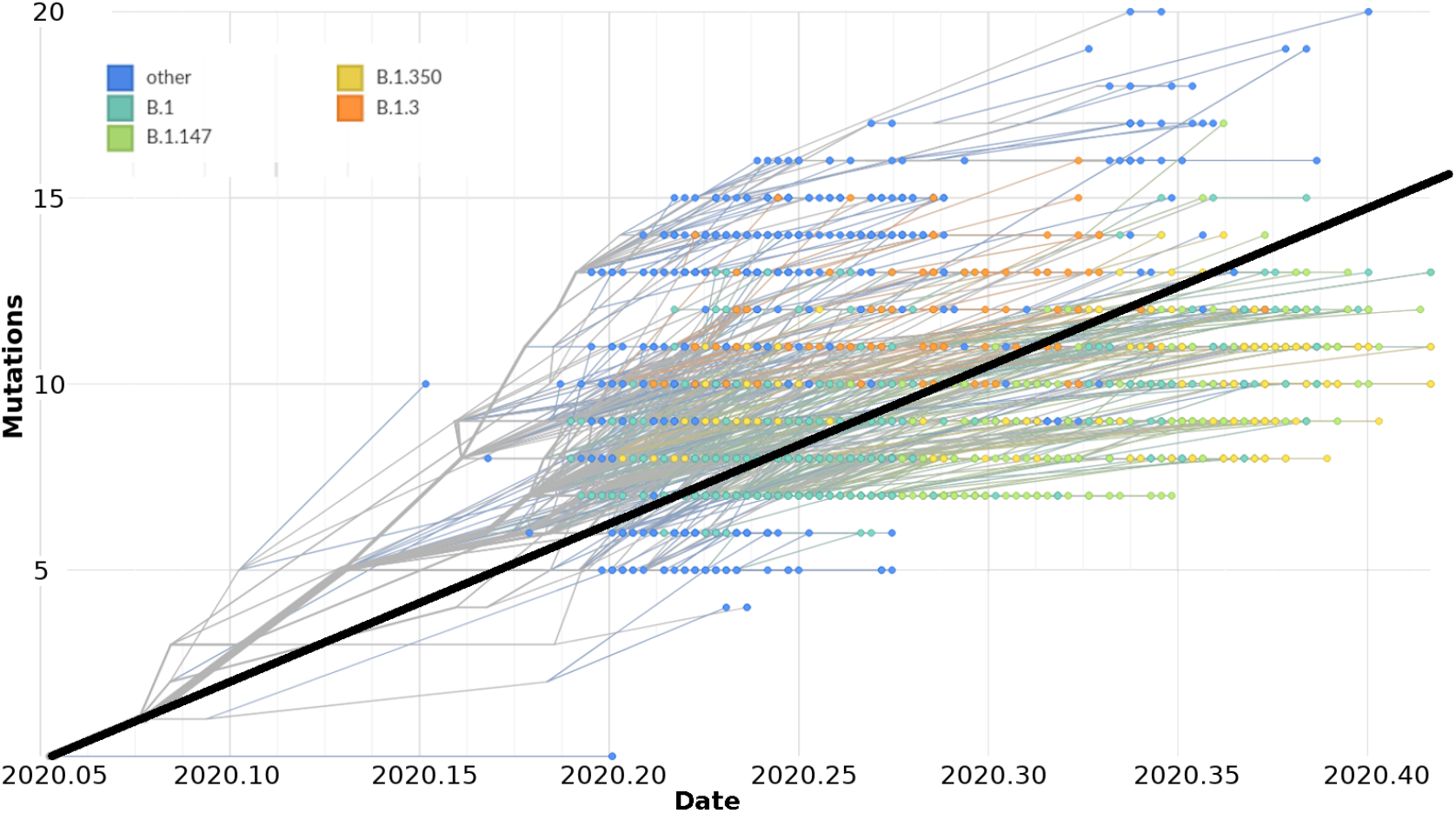
Root-to-tip regression analysis of Québec SARS-CoV-2 genomes up to June 1st, 2020. Most successful viral lineages are annotated by colors. This dataset exhibits a weak association between genetic distances and sampling dates (correlation coefficient, r = 0.33, R^2^ = 0.11, slope = 5.65×10^−4^ substitutions/site/year, *p* < 0.0001), calculated with TempEst (Rambaut et al. 2016) using the maximum-likelihood tree displayed in Fig. S4 B.

**Fig. S6.**
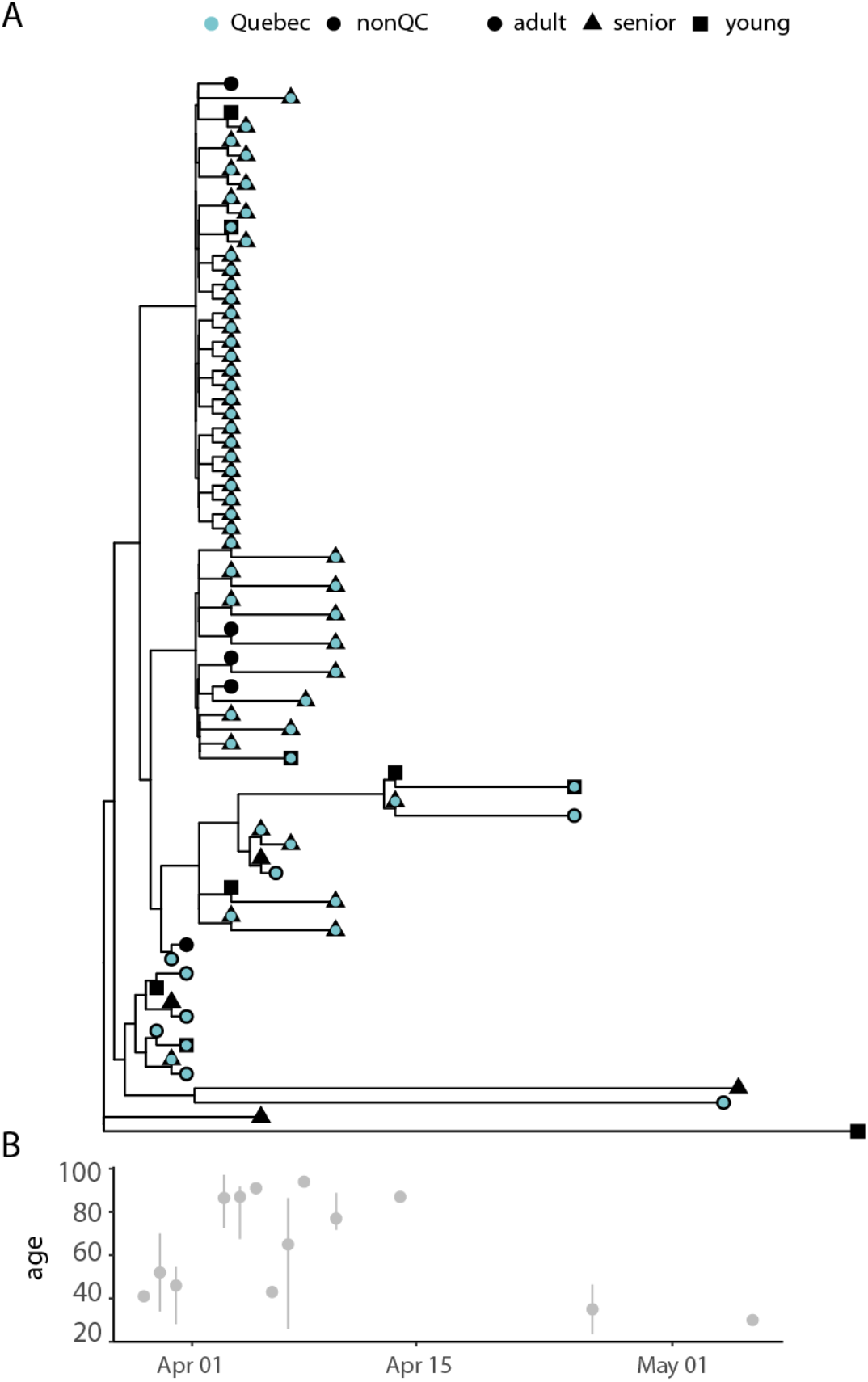
(A) Subtree of the time-scaled global phylogeny that captures a transmission lineage of B.1 that spread through a Laval long-term care facility. Black: consensus sequences from GISAID, not from Québec, Blue: Québec viral sequences. Young: < 30 years old, Adult: 30 −60 years old, Senior: > 60 years old. (B) Age distribution of the Québec cases in this transmission lineage, median age (range).

**Fig S7.**
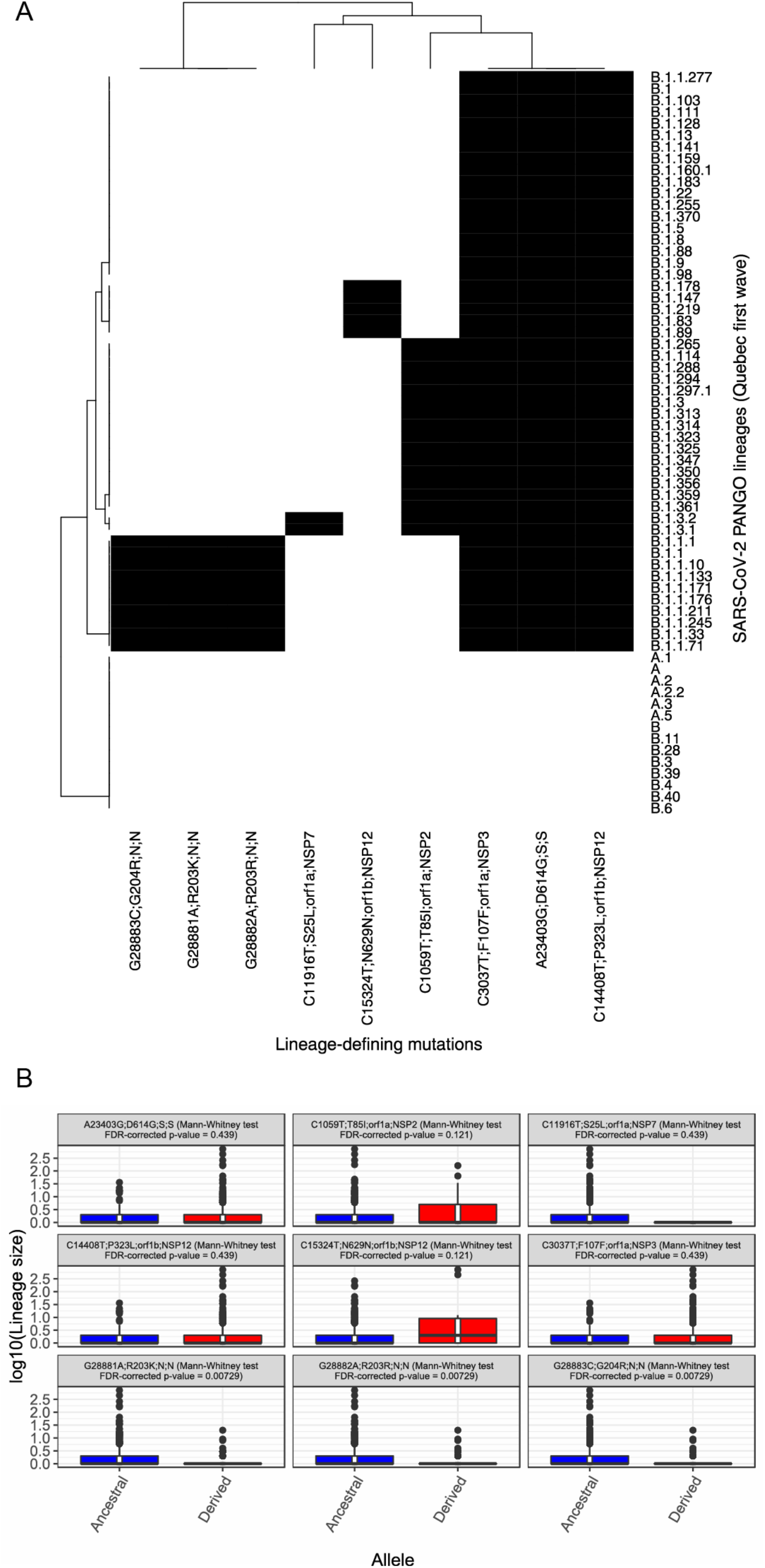
Associations between lineage-defining mutations and transmission lineage size. (A) Lineage-defining mutations. For each SARS-CoV-2 Pangolin viral lineage observed in Québec during the first wave, the nine mutations (columns) that are present in the consensus sequences of all the instances of the lineage (rows) are represented in black. The resulting heatmap was ordered by hierarchical clustering. (B) Associations between lineage-defining mutations and transmission lineage size. Each of the nine lineage-defining mutations was tested for association with transmission lineage size by comparing the transmission lineages size with the ancestral allele (blue) to the derived allele (red) (Mann-Whitney test, *P*-values reported after false-discovery rate correction for multiple hypothesis testing). The lineage-defining mutations are labeled in the following format: Ancestral nucleotide allele, Genome position, Derived nucleotide allele; Ancestral Amino Acid allele, Position In Protein, Derived Amino Acid allele; ORF; Gene.

